# Tongue Swab Xpert MTB/RIF Ultra Testing for Tuberculosis Using a Revised Consensus Protocol: A multi-country diagnostic accuracy study

**DOI:** 10.1101/2025.07.08.25330424

**Authors:** Bukola Ajide, Caitlin A. Moe, Jheannie Barrameda, Masuzyo Chirwa, Loren Rockman, Petra de Haas, Margaretha de Vos, Midori Kato-Maeda, Bianca Tasca, John Bimba, Charles Yu, Claudia M. Denkinger, Kristin Kremer, Payam Nahid, Adithya Cattamanchi, Grant Theron, Monde Muyoyeta, the R2D2 TB Network

## Abstract

**Background:** Tongue swabs are a promising specimen for tuberculosis (TB) diagnosis. In a previous study using a consensus protocol, tongue swabs tested with Xpert MTB/RIF Ultra (Xpert Ultra, Cepheid, USA) outperformed sputum smear microscopy, but a substantial proportion (6.1%) of results were non-actionable (e.g., invalid/error). We evaluated a revised protocol in four high TB burden countries.

**Methods:** Participants aged ≥12 years with presumptive TB were enrolled from outpatient clinics in the Philippines, South Africa, Nigeria, and Zambia. Tongue swabs were processed using Sample Reagent (SR, Cepheid, USA) diluted 2:1 with phosphate buffer or phosphate-buffered saline and tested with Xpert Ultra. Diagnostic performance was assessed against a culture-based microbiological reference standard and compared to sputum-based tests.

**Results:** From March to November 2024, 1168 participants were enrolled (median age 37 [IQR 28– 48] years; 46.7% female, 21.8% living with HIV, 18.5% culture-confirmed TB). The proportion of non-actionable results was 5.6% overall, but was less than 4% in all countries except South Africa (15.4%). Tongue swab sensitivity was 66.0% (95% CI 59.0–72.5); specificity was 99.6% (95% CI98.9–99.9).

**Conclusion:** The revised protocol yielded low error rates at most sites and moderate sensitivity, supporting tongue swabs as an alternative when sputum is unavailable.

## INTRODUCTION

Diagnosing tuberculosis (TB) remains a significant challenge, with approximately 25% of people with TB not diagnosed or reported to public health authorities.^1^ Among those diagnosed, fewer than half receive a World Health Organization (WHO)-recommended rapid diagnostic (WRD).^1,2^ The reliance on sputum as the primary sample type for TB diagnosis is a major barrier, particularly affecting vulnerable populations such as children, people living with HIV (PLHIV) and people with minimal symptoms.^3^ The persistent diagnostic gap underscores the urgent need for alternative approaches to enhance timely and accurate TB diagnosis, which is critical for improving clinical outcomes and curbing transmission.^4^

Tongue swabs represent a promising alternative sample type for TB diagnosis, offering several key advantages over conventional sputum samples. Swabbing the tongue surface is easier, safer and more discreet compared to sputum collection,^5^ making it a more comfortable and non-invasive option preferred by many individuals with TB.^6^ Furthermore, tongue swabs can easily be collected from people of all ages and regardless of TB symptom burden.

We and others have previously demonstrated the potential of tongue swabs for TB diagnosis when tested with Xpert MTB/RIF Ultra (Xpert Ultra, Cepheid, USA). However, variability in swab processing and collection protocols has hindered the comparability of results across different studies.^7^ To address this, we developed a consensus protocol for tongue swab Xpert Ultra testing, which was evaluated in a large-scale, multi-country study.^8^ This study reported moderate sensitivity (65.6% overall; range 37-77% across study sites) and high specificity. However, the proportion of non-actionable test results was notably high (6.1% overall; range 0-16% across study sites; >5% in 4 of 10 study sites), primarily due to cartridge pressure issues.

Subsequent research indicated that processing tongue swabs with diluted Sample Reagent (SR, Cepheid, USA) buffer enhanced liquefaction without compromising test performance.^9^ Based on these findings, we revised the consensus protocol to replace heat inactivation in TE buffer to inactivation using diluted SR buffer. In this manuscript, we report on the performance of the revised consensus protocol for Xpert Ultra testing of tongue swabs in four high TB burden countries.

## METHODS

### Study Design and Setting

We conducted a prospective, multicenter diagnostic accuracy evaluation in South Africa, Nigeria, Philippines, and Zambia as part of the Rapid Research in Diagnostics Development for TB Network (R2D2 TB Network) and Assessing Diagnostics At Point-of-care for Tuberculosis (ADAPT) studies. Primary objectives were to evaluate the diagnostic accuracy of a revised consensus protocol of tongue swab Xpert Ultra testing to detect pulmonary TB among adolescents and adults. Secondary objectives were to quantify the proportion of non-actionable results (*e*.*g*., invalid or error) and compare to that of conventional sputum-based TB tests (smear microscopy and Xpert Ultra).

### Study Participants

We screened consecutive people aged 12 years of age or older presenting to outpatient health centers for presumptive TB as part of the R2D2 TB Network (South Africa) and ADAPT (Nigeria, Philippines, Zambia) studies. We included people who reported a cough of at least 2 weeks or had a TB risk factor (HIV-positive, TB contact, or mining history) plus a positive TB screening test (abnormal chest X-ray or, for PLHIV, C-reactive protein [CRP]>5mg/dL). People were excluded if they had taken preventive or active TB treatment in the last 12 months, had taken medication with anti-mycobacterial activity in the last 2 weeks, or were unwilling or unable to provide informed consent.

### Study Procedures

Detailed study procedures have been reported elsewhere.^8,10,11^ Briefly, demographic and medical history data were collected using standardized case report forms. Two to three sputum samples were collected from each participant for reference standard testing. Blood was collected for HbA1c, HIV testing and CD4 count (if HIV-positive). Tongue swabs (Copan FLOQswab 520CS01) were collected into cryovial tubes prior to sputum collection for Xpert Ultra testing. Participants were requested to not eat or drink for at least 30 minutes prior to tongue swab collection. Using a back-front and left-right motion, the tongue was swabbed from the back of the top of the tongue and as far back as possible without creating a gag reflex (about ¾ of the visible tongue dorsum) for 30 seconds. Immediately after collection, the tongue swab head was snapped off into the dry cryovial tube. Tubes were closed and transported to a research laboratory for testing within 24 hours of collection.

### Tongue Swab Xpert Ultra

Tongue swabs collected into dry tubes were processed for Xpert Ultra testing using the revised consensus protocol.^12^ The revision replaced TE buffer plus heat inactivation with Xpert Sample Reagent (SR) diluted 2:1 with phosphate buffer (PB) or phosphate buffered saline (PBS) depending on local availability.^13^ After incubating swabs for 15 minutes in 700 µL of diluted SR buffer, 1.5 mL of the solution was added to the Xpert Ultra cartridge. Xpert Ultra testing then proceeded as per manufacturer instructions. Results were recorded as positive for *M. tuberculosis* (any semi-quantitative grade including ‘Trace’), negative for *M. tuberculosis*, or non-actionable (*i*.*e*., invalid, error, or no result).

### Sputum Comparator Tests

Tongue swab Xpert Ultra results were compared to sputum smear microscopy and sputum Xpert Ultra results. LED fluorescence microscopy with auramine staining (two replicates) was performed on decontaminated sputum specimens following WHO-recommended protocols.^14^ Results were recorded as smear-positive if one or more acid-fast bacilli (AFB) were seen on either smear, or as negative if no AFB were seen on both smears.

Xpert Ultra testing was performed on the remainder of the first sputum specimen in accordance with manufacturer recommendations. Results were recorded as positive for *M. tuberculosis* (semi-quantitative grade of ‘Very Low’ or higher), negative for *M. tuberculosis* or non-actionable (*i*.*e*., invalid, error, or no result).

### Reference Standard

The primary microbiological reference standard (MRS) was based on the results of liquid media culture using Mycobacterium Growth Indicator Tubes (MGIT, Becton Dickinson, Franklin Lakes, NJ, USA). Participants were classified as TB-positive if at least one of two sputum cultures tested positive for *M. tuberculosis* complex (MTBC), and as TB-negative if both sputum cultures were negative for MTBC. Participants with contaminated results for one or both cultures and no positive culture results were classified as indeterminate. Laboratory personnel performing culture testing were blinded to the results of the index and comparator tests.

### Outcome Measures

The primary outcome was the diagnostic accuracy of tongue swab Xpert Ultra, as measured by its sensitivity and specificity in comparison to the MRS. Secondary outcomes included the proportion of non-actionable results generated by tongue swab Xpert Ultra testing and the sensitivity and specificity differences between tongue swab Xpert Ultra and sputum-based comparator tests.

### Statistical Analysis

Proportions of non-actionable results from Xpert Ultra were estimated as simple proportions with 95% confidence intervals. Sensitivity and specificity of tongue swabs were calculated against the MRS and reported with exact 95% confidence intervals, both overall and within key subgroups (country, sex, HIV status, diabetes, AFB smear status, and sputum collection method). We compared differences in sensitivity and specificity between tongue swab Xpert Ultra and sputum-based comparator tests using McNemar’s test for paired proportions. Analyses were conducted in Stata version 18 (StataCorp, USA).

### Ethics Statement

This study was registered on ClinicalTrials.gov (NCT04923958 and NCT05941052). Written informed consent was obtained from all study participants. Ethical approval for the study was obtained from institutional review boards and/or research ethics committees at the University of California San Francisco (USA), University of Heidelberg (Germany), Abuja Health Research Ethics Committee (Nigeria), Stellenbosch University (South Africa), De la Salle Medical and Health Sciences Institute (Philippines), and the University of Zambia (Zambia).

This manuscript adheres to the STARD guidelines for reporting diagnostic accuracy studies.^15^

## RESULTS

### Study Population

Between March and November 2024, 1281 people with presumptive TB were screened across all study sites and 75 (5.9%) were excluded (**Figure 1**). Of the remaining 1206 participants enrolled, 38 (3.2%) with missing or improperly processed tongue swab Xpert Ultra results were excluded from all analyses.

**Figure 1.**
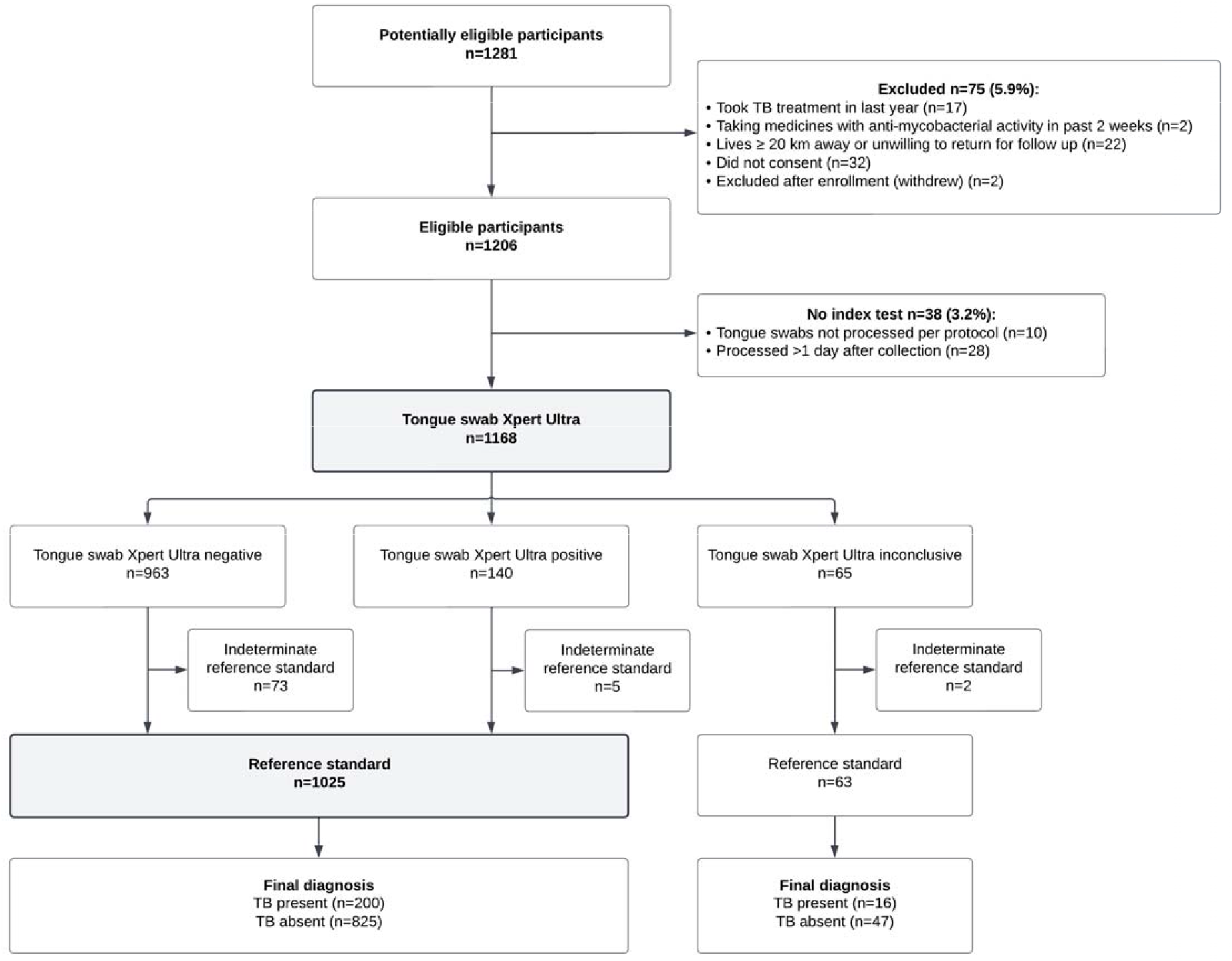
Participant flow diagram.

The overall median age of the 1168 participants included was 37 years (interquartile range [IQR]: 28-48 years), 545 (46.7%) were female, 254 (21.8%) were PLHIV and 254 (21.8%) reported a prior history of TB (**Table 1**). Most (95.2%) participants reported a cough of 2 weeks or more, while 56 (4.8%) were enrolled based on having a TB risk factor and positive screening test (abnormal chest X-ray and/or CRP> 5mg/dL). Sputum induction was required for 113 (9.7%) participants unable to spontaneously expectorate sputum for TB testing. The prevalence of culture-positive TB was 18.5% (216/1168).

**Table 1.** Participant characteristics.

|  | Overall<br>(N=1168) | Philippines<br>(N=295) | South Africa<br>(N=272) | Zambia<br>(N=326) | Nigeria<br>(N=275) |
| --- | --- | --- | --- | --- | --- |
| Age, median (IQR) | 37 (28, 48) | 40 (27, 55) | 37 (29, 47) | 37 (28, 47) | 36 (26, 46) |
| Female sex <sup>a</sup> | 545 (46.7%) | 140 (47.5%) | 158 (58.1%) | 118 (36.2%) | 129 (46.9%) |
| *HIV status* <sup>b</sup> |  |  |  |  |  |
| Negative | 912 (78.2%) | 291 (98.6%) | 181 (66.5%) | 211 (65.1%) | 229 (83.3%) |
| Positive | 254 (21.8%) | 4 (1.4%) | 91 (33.5%) | 113 (34.9%) | 46 (16.7%) |
| History of TB | 254 (21.8%) | 81 (27.5%) | 70 (25.7%) | 78 (23.9%) | 25 (9.1%) |
| Cough ≥ 2 weeks | 1112 (95.2%) | 292 (99.0%) | 245 (90.1%) | 311 (95.4%) | 264 (96.0%) |
| Symptomatic <sup>c</sup> | 1150 (98.5%) | 295 (100%) | 257 (94.5%) | 326 (100%) | 272 (98.9%) |
| Sputum induced | 113 (9.7%) | 101 (34.2%) | 12 (4.4%) | 0 (0%) | 0 (0%) |
| AFB smear-positive | 146 (12.5%) | 39 (13.2%) | 14 (5.2%) | 43 (13.2%) | 50 (18.2%) |
| *Microbiological reference standard* |  |  |  |  |  |
| TB Negative | 872 (74.7%) | 214 (72.5%) | 221 (81.3%) | 230 (70.6%) | 207 (75.3%) |
| TB Positive | 216 (18.5%) | 67 (22.7%) | 43 (15.8%) | 52 (16.0%) | 54 (19.6%) |
| Indeterminate | 80 (6.8%) | 14 (4.8%) | 8 (2.9%) | 44 (13.5%) | 14 (5.1%) |
| *Sputum Xpert Ultra* <sup>d</sup> |  |  |  |  |  |
| Negative | 950 (82.3%) | 237 (80.6%) | 224 (83.9%) | 275 (85.4%) | 214 (78.7%) |
| Non-actionable (error/invalid/no result) | 15 (1.3%) | 6 (2.0%) | 5 (1.9%) | 0 (0.0%) | 4 (1.5%) |
| Positive | 190 (16.5%) | 51 (17.3%) | 38 (14.2%) | 47 (14.6%) | 54 (19.9%) |
| Very Low | 18 (9.5%) | 6 (11.8%) | 5 (13.2%) | 3 (6.4%) | 4 (7.4%) |
| Low | 40 (21.1%) | 16 (31.4%) | 7 (18.4%) | 9 (19.2%) | 8 (14.8%) |
| Medium | 49 (25.8%) | 12 (23.5%) | 8 (21.1%) | 13 (27.7%) | 16 (29.6%) |
| High | 83 (43.7%) | 17 (33.3%) | 18 (47.4%) | 22 (46.8%) | 26 (48.2%) |

Abbreviations: IQR, interquartile range; TB, tuberculosis; AFB, acid-fast bacillus
<sup>a</sup> excluding n=2 participants who declined to state
<sup>b</sup> missing for n=2 participants
<sup>c</sup> any cough, fever, night sweats or weight loss
<sup>d</sup> excluding n=13 participants with trace sputum Xpert Ultra result

### Performance of Tongue Swab Xpert Ultra

The proportion on non-actionable tongue swab Xpert Ultra results was 5.6% (65/1168) overall.This proportion was below 4% in three of four countries (4/275 in Nigeria, 11/295 in the Philippines and 8/326 in Zambia) and was highest in South Africa at 15.4% (42/272) (**Table S1**). There was no clear association between time and frequency of these non-actionable tongue swab Xpert Ultra results (**Figure S1**). No demographic or clinical characteristics were associated with non-actionable tongue swab Xpert Ultra results (**Table S2**).

In contrast, the proportion of non-actionable sputum Xpert Ultra results was 1.3% (15/1168) overall and similar across countries (**Table 1**). No participant had a non-actionable result on both tongue swab and sputum Xpert Ultra testing.

The overall sensitivity of tongue swab Xpert Ultra against the MRS was 66.0% (95% CI 59.0-72.5), and specificity was 99.6% (95% CI 98.9-99.9) (**Table 2**). Sensitivity ranged by country from 47.6% (95% CI 34.9-60.6) in the Philippines to 79.4% (95% CI 62.1-91.3) in South Africa (**Table 2**). Among participants unable to spontaneously produce sputum and requiring induction, tongue swab Xpert Ultra had a sensitivity of 42.1% (95% CI, 20.3-66.5) and a specificity of 100% (95% CI, 95.7-100). Among PLHIV, the sensitivity of tongue swabs was 60.6% (95% CI 42.1-77.1) and specificity was 100% (98.0-100).

**Table 2.** Sensitivity and specificity of tongue swab Xpert Ultra overall and by subgroup (N=1025)^a^.

|  | Microbiological reference standard (MRS) |  |
| --- | --- | --- |
|  | Sensitivity, % (95%CI)<br>[n/N] | Specificity, % (95%CI)<br>[n/N] |
| **Overall** | 66.0 (59.0, 72.5) [132/200] | 99.6 (98.9, 99.9) [822/825] |
| **Country** |  |  |
| Philippines | 47.6 (34.9, 60.6) [30/63] | 100 (98.2, 100) [207/207] |
| South Africa | 79.4 (62.1, 91.3) [27/34] | 100 (98.1, 100) [190/190] |
| Zambia | 66.0 (51.2, 78.8) [33/50] | 99.1 (96.8, 99.9) [222/224] |
| Nigeria | 79.2 (65.9, 89.2) [42/53] | 99.5 (97.3, 100) [203/204] |
| **HIV Status<sup>b</sup>** |  |  |
| Positive | 60.6 (42.1, 77.1) [20/33] | 100 (98.0, 100) [182/182] |
| Negative | 67.1 (59.4, 74.1) [112/167] | 99.5 (98.6, 99.9) [638/641] |
| **Sex** |  |  |
| Female | 68.6 (54.1, 80.9) [35/51] | 100 (99.1, 100) [426/426] |
| Male | 65.1 (56.9, 72.7) [97/149] | 99.2 (97.8, 99.8) [396/399] |
| **Diabetes** |  |  |
| No | 65.1 (57.5, 72.2) [112/172] | 99.6 (98.9, 99.9) [760/763] |
| Yes | 71.4 (51.3, 86.8) [20/28] | 100 (94.2, 100) [62/62] |
| **Current smoker** |  |  |
| No | 64.8 (56.3, 72.6) [92/142] | 99.5 (98.5, 99.9) [598/601] |
| Yes | 69.0 (55.5, 80.5) [40/58] | 100 (98.4, 100) [224/224] |
| **AFB smear status** |  |  |
| Negative | 26.9 (16.8, 39.1) [18/67] | -- |
| Positive | 85.7 (78.6, 91.2) [114/133] | -- |
| **Sputum collection** |  |  |
| Expectorated | 68.5 (61.2, 75.2) [124/181] | 99.6 (98.8, 99.9) [739/742] |
| Induced | 42.1 (20.3, 66.5) [8/19] | 100 (95.7, 100) [83/83] |

<sup>a</sup> excludes n=65 participants with non-actionable Xpert Ultra result and n=78 with indeterminate MRS result
<sup>b</sup> HIV status missing for n=2 participants
Abbreviations: CI, confidence interval, AFB, acid fast bacillus

### Comparison to Sputum-Based Index Tests

Positive concordance between tongue swab Xpert Ultra and sputum Xpert Ultra was 76.0% overall and correlated with sputum Xpert Ultra semi-quantitative grade: Positive concordance of tongue swab Xpert Ultra results ranged from 12.5% (2/16) among participants with “Very Low” positive sputum Xpert Ultra results to 94.7% (71/75) among those with “High” positive sputum Xpert Ultra results (**Table 3**).

**Table 3.** Concordance between tongue swab Xpert Ultra and sputum Xpert Ultra (N=1076) ^a^.

| Sputum Xpert Ultra | Tongue Swab Xpert Ultra<br>% [n/N] |
| --- | --- |
| **Positive concordance** | 76.0 [133/175] |
| High | 94.7 [71/75] |
| Medium | 88.9 [40/45] |
| Low | 51.3 [20/39] |
| Very low | 12.5 [2/16] |
| **Negative concordance** | 99.7 [898/901] |

<sup>a</sup> Note. Excludes n=65 participants with non-actionable tongue swab Xpert Ultra result, n=15 with non-actionable sputum Xpert Ultra result, and n=12 with Trace sputum Xpert Ultra result

In comparison to sputum smear microscopy, tongue swab Xpert Ultra had similar sensitivity (66.0% vs. 66.5%, difference −0.5%, 95% CI −7.0 to +6.0, p>0.99) and specificity (99.6% vs. 99.8%, difference −0.1%, 95% CI −0.7 to +0.4, p>0.99) (**Table 4**). In comparison to sputum Xpert Ultra, sensitivity was lower (68.4% vs. 87.2%, difference −18.7%, 95% CI −25.0 to −12.4, p>0.01) but specificity was similar (99.6% vs. 99.1%, difference 0.5%, 95% CI −0.3 to +1.3, p=0.29). In a secondary analysis where “Trace” sputum Xpert Ultra results were considered positive, we similarly found that the sensitivity of tongue swab Xpert Ultra was lower (66.7% vs. 87.7%, difference −21.0%, 95% CI −27.4 to −14.6, p>0.01) than sputum Xpert Ultra, but specificity was similar (99.6% vs. 98.8%, difference 0.9%, 95% CI −0.06 to +1.8, p=0.07) (**Table S3**).

**Table 4.** Diagnostic performance of tongue swab Xpert Ultra compared to sputum-based TB tests.

|  | Sensitivity |  |  | Specificity |  |  |
| --- | --- | --- | --- | --- | --- | --- |
|  | % (95%CI) [n/N] | Difference, %<br>(95% CI) | p-value | % (95%CI) [n/N] | Difference, %<br>(95% CI) | p-value |
| Tongue swab Xpert Ultra vs. sputum smear microscopy (N=1025)<sup>a</sup> |  |  |  |  |  |  |
| Tongue swab Xpert Ultra | 66.0 (59.0, 72.5) [132/200] | -0.5 | >0.99 | 99.6 (98.9, 99.9) [822/825] | -0.1 | >0.99 |
| Smear microscopy | 66.5 (59.5, 73.0) [133/200] | (-7.0, +6.0) |  | 99.8 (99.1, 100) [823/825] | (-0.7, +0.4) |  |
| Tongue swab Xpert Ultra vs sputum Xpert Ultra (N=1001)<sup>b</sup> |  |  |  |  |  |  |
| Tongue swab Xpert Ultra | 68.4 (61.3, 75.0) [128/187] | -18.7 | <0.001 | 99.6 (98.9, 99.9) [811/814] | +0.5 | 0.29 |
| Sputum Xpert Ultra | 87.2 (81.5, 91.6) [163/187] | (-25.0, -12.4) |  | 99.1 (98.2, 99.7) [807/814] | (-0.3, +1.3) |  |

<sup>a</sup> Excludes n=80 participants with indeterminate MRS result and n=63 with non-actionable tongue swab Xpert Ultra result
<sup>b</sup> Excludes n=80 participants with indeterminate MRS result, n=63 with non-actionable tongue swab Xpert Ultra result, n=13 with non-actionable sputum Xpert Ultra result, and n=11 with Trace sputum Xpert Ultra result
Abbreviations: TB, tuberculosis; CI, confidence interval

## DISCUSSION

In this multi-country assessment of the diagnostic accuracy of tongue swab Xpert Ultra testing with a revised sample processing method incorporating diluted SR buffer, we found sensitivity to be modest at 66%, which is comparable to sputum smear microscopy. Importantly, tongue swab Xpert Ultra testing was positive in 38% of people with TB who were unable to expectorate sputum, highlighting its potential utility in this subgroup. Although the overall proportion of non-actionable results remained high at 5.6% (65/1168), this was primarily driven by one site (South Africa, 15.4%). Taken together, these findings suggest that while tongue swab Xpert Ultra testing may have limited broad application, it could be a valuable alternative for people with presumptive TB who are unable to produce sputum.

The original consensus protocol for tongue swab Xpert Ultra testing used heat inactivation in TE buffer instead of inactivation with SR because previous studies indicated that the use of SR buffer may worsen analytical sensitivity.^9,16^ Similar to a recent study using banked tongue swab samples,^17^ our findings suggest that the use of diluted SR buffer does not compromise the clinical sensitivity of tongue swab Xpert Ultra testing. Sensitivity was similar to what we observed with the original consensus protocol, providing further evidence that sensitivity is below the minimum target recommended in the target product profile for a low complexity, non-sputum based rapid diagnostic test.^18^

The correlation between tongue swab Xpert Ultra sensitivity and sputum Xpert Ultra semi-quantitative grades was also evident in our study, with notably higher sensitivity among samples with “High” sputum grades (95.8%) compared to those with “Very Low” (8.3%) grades. This finding aligns with other studies,^8,16^ reinforcing the notion that tongue swabs may be less effective in detecting lower bacterial loads. The high specificity (>99%) observed in our study also aligns with previous research,^7^ indicating that tongue swabs are highly reliable when results are positive.

The re-introduction of SR buffer at a 2:1 dilution mostly resolved the cartridge pressure issues and related high error rates observed with the original consensus protocol, except at one site. The high proportion of non-actionable results observed in South Africa (42/272, 15%) was different to what was observed when using the original consensus protocol at the same study site (4/198, 2%)^8^ and in a South African study that used banked tongue swab specimens (2/110, 1.8%).^17^ The reason for the high proportion of non-actionable results in our study in South Africa is unclear. We cannot exclude the possibility of site-specific issues related to Xpert Ultra cartridge lot and GeneXpert module performance, although sputum Xpert Ultra non-actionable results were uncommon. It is also possible that the presence of PCR inhibitors in the tongue swab sample matrix vary by population, but PCR inhibition was not observed at the same site in South Africa when using the original consensus protocol.

The strengths of our study include the implementation of a standardized testing protocol across diverse high TB burden settings and a robust sample size. However, key limitations include the lack of a direct head-to-head comparison with previous methodologies for tongue swab Xpert Ultra testing and a clearly identifiable reason for the high proportion of non-actionable results in South Africa. As such, the revised consensus protocol described here should be piloted before use in specific settings.

## Conclusions

In conclusion, a revised consensus methodology for testing tongue swabs with Xpert Ultra showed similar diagnostic accuracy as reported in previous studies and overall resulted in most sites having <4% non-actionable results. Future studies should focus on further improving sample collection and processing protocols, and on comparison to simpler near point-of-care technologies that have been designed specifically for swab-based molecular testing.^11^

## Data Availability

All data produced in the present study are available upon reasonable request to the authors

## Acknowledgments

We gratefully acknowledge the individuals who participated in this study and extend our sincere thanks to the administrative teams and clinical staff at the participating health centers for their support and dedication. This work would not have been possible without their partnership and collaboration.

## SUPPLEMENTAL TABLES

**Table S1.** Tongue swab Xpert Ultra Results by country (N=1168).

|  | Overall<br>(N=1168) | Philippines<br>(N=295) | South Africa<br>(N=272) | Zambia<br>(N=326) | Nigeria<br>(N=275) |
| --- | --- | --- | --- | --- | --- |
| Negative | 963 (82.5%) | 254 (86.1%) | 203 (74.6%) | 281 (86.2%) | 225 (81.8%) |
| Non-actionable (error/invalid/no result) | 65 (5.6%) | 11 (3.7%) | 42 (15.4%) | 8 (2.5%) | 4 (1.5%) |
| Positive | 140 (12.0%) | 30 (10.2%) | 27 (9.9%) | 37 (11.4%) | 46 (16.7%) |
| High | 0 (0%) | 0 (0%) | 0 (0%) | 0 (0%) | 0 (0%) |
| Medium | 6 (4.3%) | 0 (0%) | 1 (3.7%) | 3 (8.1%) | 2 (4.4%) |
| Low | 81 (57.9%) | 15 (50.0%) | 17 (63.0%) | 20 (54.1%) | 29 (63.0%) |
| Very Low | 35 (25.0%) | 7 (23.3%) | 5 (18.5%) | 11 (29.7%) | 12 (26.1%) |
| Trace | 18 (12.9%) | 8 (26.7%) | 4 (14.8%) | 3 (8.1%) | 3 (6.5%) |

**Figure S1.**
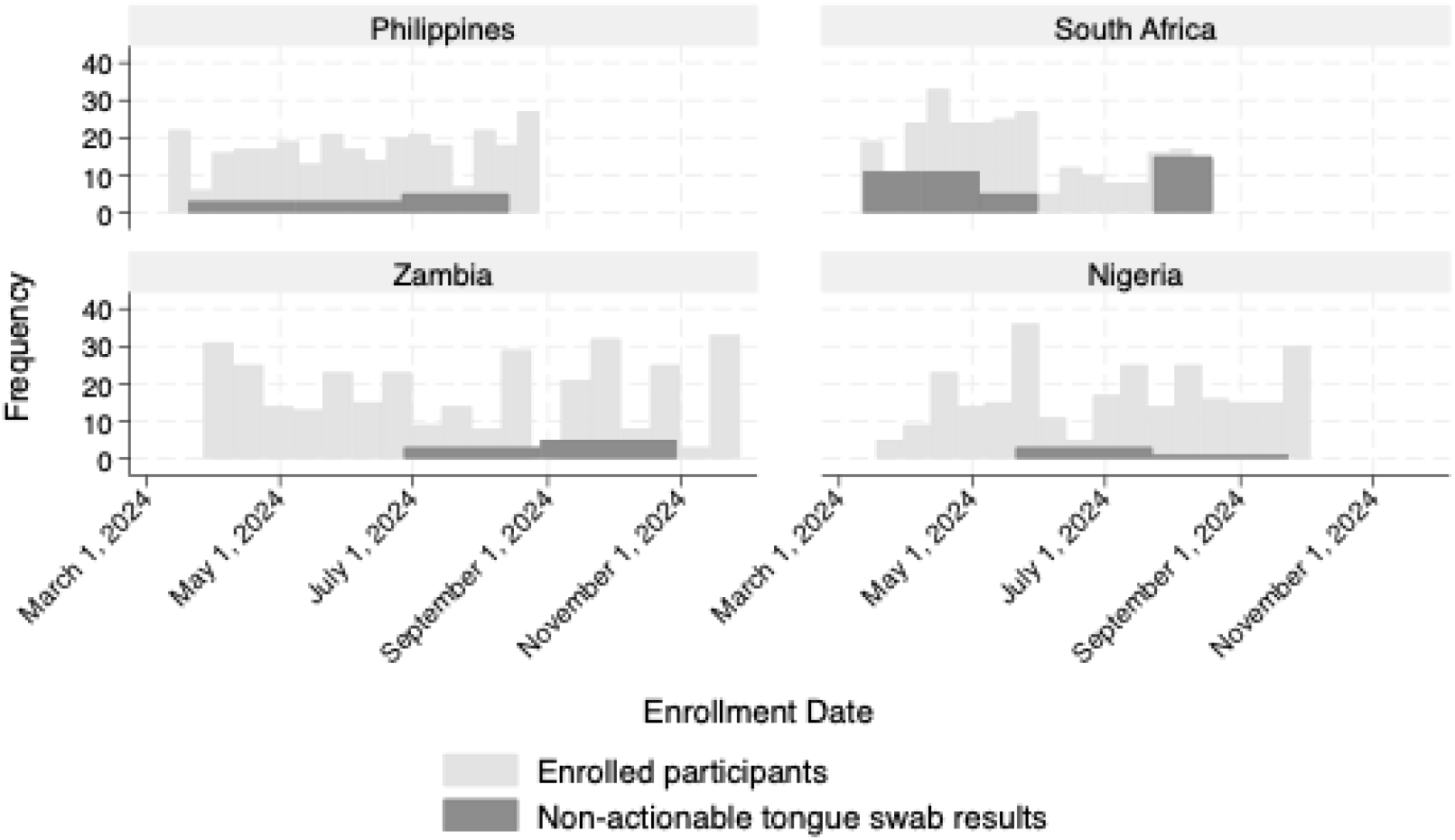
Non-actionable tongue swab Xpert Ultra results over time, by enrollment site.

**Table S2.** Characteristics associated with non-actionable tongue swab Xpert Ultra results (N=1165)^a^.

|  | OR (95% CI) | p-value |
| --- | --- | --- |
| Age, years | 1.01 (0.99, 1.03) | 0.28 |
| Enrollment country |  |  |
| Nigeria | Ref |  |
| Philippines | 2.70 (0.84, 8.67) | 0.095 |
| South Africa | 11.39 (4.00, 32.45) | <0.001 |
| Zambia | 1.59 (0.47, 5.40) | 0.46 |
| Sex |  |  |
| Male | Ref |  |
| Female | 1.16 (0.68, 1.96) | 0.59 |
| HIV Status |  |  |
| Negative | Ref |  |
| Positive | 1.46 (0.81, 2.62) | 0.21 |

<sup>a</sup> Excludes n=2 with missing HIV status
Abbreviations: CI, confidence interval; OR, odds ratio

**Table S3.**
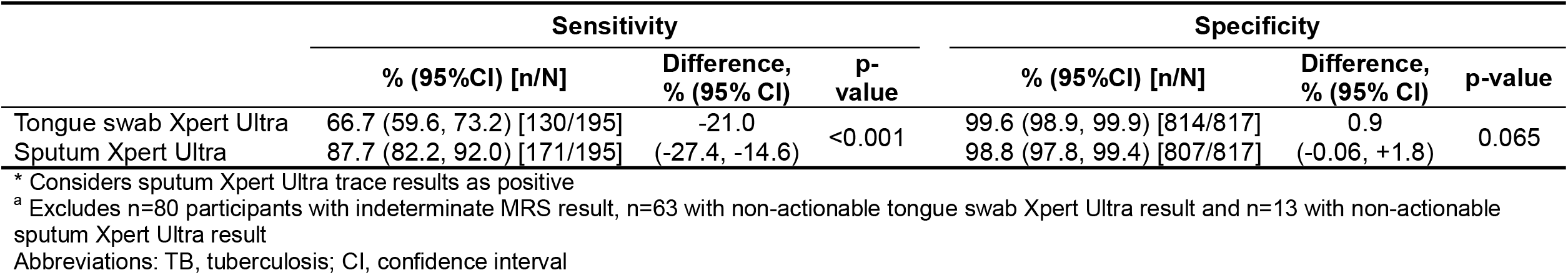
Diagnostic performance of tongue swab Xpert Ultra compared to sputum Xpert Ultra: Secondary analysis* (N=1012)^a^.

|  | Sensitivity |  |  | Specificity |  |  |
| --- | --- | --- | --- | --- | --- | --- |
|  | % (95%CI) [n/N] | Difference, % (95% CI) | p-value | % (95%CI) [n/N] | Difference, % (95% CI) | p-value |
| Tongue swab Xpert Ultra | 66.7 (59.6, 73.2) [130/195] | -21.0 | <0.001 | 99.6 (98.9, 99.9) [814/817] | 0.9 | 0.065 |
| Sputum Xpert Ultra | 87.7 (82.2, 92.0) [171/195] | (-27.4, -14.6) |  | 98.8 (97.8, 99.4) [807/817] | (-0.06, +1.8) |  |

\* Considers sputum Xpert Ultra trace results as positive
<sup>a</sup> Excludes n=80 participants with indeterminate MRS result, n=63 with non-actionable tongue swab Xpert Ultra result and n=13 with non-actionable sputum Xpert Ultra result
Abbreviations: TB, tuberculosis; CI, confidence interval

